# PanCNV-Explorer: A pan-cancer resource to analyze copy number variations

**DOI:** 10.1101/2024.09.23.24314206

**Authors:** Kevin Kornrumpf, Jürgen Dönitz

## Abstract

**Introduction:** Copy number variations (CNVs) are structural genomic alterations that involve changes in the number of copies of specific DNA regions. These variations can include deletions, duplications, and more complex rearrangements, and play a critical role in cancer progression by amplifying oncogenes, deleting tumor suppressor genes, or altering other key genomic regions. Despite the importance of CNVs in cancer biology, there is a lack of comprehensive resources that aggregate CNV data across multiple cancer types, impeding the exploration of their role in different malignancies. The PanCNV-Explorer was developed to fill this gap by providing a pan-cancer resource that integrates CNV data for over 21 cancer types.

**Methods:** The PanCNV-Explorer was developed, utilizing data from repositories such as COSMIC, DepMap, DGV, and dbVAR. Amplifications and deletions can be visualized, with detailed chromosome-specific plots highlighting CNV patterns. In addition, a genome browser has been implemented to display regions of interest for searching for genes of potential relevance.

**Results:** The database contains over 5 million CNVs from 15,809 samples, spanning 21 primary cancer types. Pathogenic CNVs from ClinVar and DepMap were compared to benign data from DGV and dbVAR, allowing a comprehensive analysis across tissues. CNV patterns were visualized for individual cancers. We demonstrate with use cases, focusing on common oncogenes such as MYC and tumor suppressor genes such as TP53, the ability of the resource to search for cancer driver genes. For example, chromosome 12 in pancreatic cancer showed frequent amplifications in the region containing the KRAS oncogene. In another example, we show how driver gene candidates already described in certain cancer entities can also be found with high frequency in other entities.

**Conclusion:** The PanCNV-Explorer provides a valuable resource for pan-cancer CNV analysis, facilitating the exploration of CNV-driven tumor heterogeneity. The integrated visualization tools and genome browser enable detailed exploration of genomic regions, enhancing cancer research. As a valuable and easy-to-use platform that allows researchers to quantify and compare CNVs across cancer types, the PanCNV-Explorer is an essential tool for advancing cancer genomics research. The main page is available at https://mtb.bioinf.med.uni-goettingen.de/pancnv-explorer/.

## Introduction

Copy number variations (CNVs) are a type of structural variation in the human genome characterized by changes in the number of copies of specific regions of the chromosomes. These variations can range from a few kilobases to several megabases in length and include deletions, duplications, or more complex rearrangements [1, 2]. CNVs are a ubiquitous feature of the human genome, contributing to genetic diversity and playing an important role in both normal development and disease pathogenesis [3].

CNVs make up a significant portion of the human genome, accounting for approximately 5-9% of the nucleotides [4]. While CNVs can be benign and contribute to phenotypic variability, they can also disrupt gene function or alter gene dosage, resulting in disease. They have been implicated in a variety of genetic disorders, including neurodevelopmental disorders, immunodeficiency, and cancer [5].

Especially in the context of cancer, CNVs are of particular interest because of their impact on large amount of genes and their high potential to drive tumorigenesis and cancer progression. They contribute to cancer development by amplifying oncogenes, deleting tumor suppressor genes, or creating fusion genes that increase malignancy [6]. For example, the amplification of the MYC oncogene or the deletion of the TP53 tumor suppressor gene are CNV events frequently observed in various types of cancer [7, 8].

In addition, CNVs may contribute to cancer progression through more complex mechanisms, such as altering gene expression in pathways that regulate the cell cycle, apoptosis, and DNA repair [9]. The heterogeneity of CNV patterns across cancer types underscores their importance as biomarkers for diagnosis and as targets for therapeutic intervention [10].

Repetitive sequences in the human genome, such as Alu elements, long interspersed nuclear elements (LINEs), and segmental duplications, are known to predispose genomic regions to structural variations. These sequences can cause non-allelic homologous recombination (NAHR) and other recombination events that can result in chromosomal rearrangements, deletions, and duplications. The instability introduced by these repetitive sequences can result in CNVs that disrupt gene function or regulatory regions, potentially initiating or promoting tumorigenesis [11].

The presence of repetitive sequences can also increase the likelihood of chromosomal damage during DNA replication and repair, further promoting the formation of CNVs [12]. This is particularly relevant in cancer cells of genomic instability, where the accumulation of CNVs can accelerate cancer progression by continuously altering the genomic landscape of the tumor [13].

The mechanisms underlying the formation of CNVs and their impact on cancer progression remain incompletely understood [10, 14]. For example, in pancreatic ductal adenocarcinoma, amplifications in genes such as KRAS, MYC, and GATA6, and deletions in TP53, CDKN2A, CDKN2B, and SMAD4 are frequently observed and serve as key indicators of tumorigenesis [15]. However, for cancer in general and many other cancer types, the available data on CNVs are sparse and remain elusive, highlighting a significant gap in the current understanding of their role in different malignancies.

On the other side, there are many CNV that are specific to cancer entities and are used to define sub-entities. In diffuse large B-cell lymphoma (DL-BCL), the activated B-cell-like (ABC) subtype is associated with recurrent genetic alterations, including deletions in *TNFAIP3* (6q23), *CDKN2A/B* (9p21) and *PRDM1* (6q21), and amplifications in *SPIB* (19q13) and *BCL2* (18q21) [16]. Similarly, recurrent duplications in *VIPR2* (7q36.3) and deletions in *NRXN1* (2p16.3) have been identified in neurodevelopmental and psychiatric disorders such as autism, schizophrenia and bipolar disorder [17].

For individual cancers, specific CNVs are well characterised, but a comprehensive pan-cancer approach is still lacking. Data resources such as COSMIC provide extensive CNV datasets, but do not provide graphical interfaces for comparing different cancer entities. A first effort to address this gap is CNVIntegrate, which compiles CNV data from multiple sources and a national study, providing an interactive platform for data exploration. However, the search is focused gene based, limiting the exploration for unknown CNV patterns [18].

A pan-cancer analysis of CNVs provides insights into both common and cancer-specific molecular aberrations, thereby advancing our understanding of cancer biology. Understanding CNV-driven tumor heterogeneity is critical because it provides insight into the mechanisms of underlying therapeutic resistance and thus contributes to the development of more effective and personalized cancer therapies.

A comprehensive approach includes the refinement of cancer classification beyond tissue origin and emphasises the importance of CNV analysis.

To address this gap, we introduce the PanCNV-Explorer, a comprehensive and integrated resource for copy number variation data. This new database combines information from established repositories such as COSMIC and dbVAR, offering extensive CNV data for over 21 cancer types, to a new pan-cancer data resource. The PanCNV-Explorer features a user-friendly web interface that facilitates easy access to individual datasets, along with advanced visualization tools and detailed data tables. An integrated genome browser further enhances the user experience by allowing detailed exploration of genomic regions frequently affected by CNVs in specific cancers. This enables users to identify and study relevant genes. The website is available at https://mtb.bioinf.med.uni-goettingen.de/pancnv-explorer/.

## Materials and methods

### Packages and frameworks

The backend of PanCNV-Explorer is developed in Python v3.9 [https://www.python.org/] using packages like Flask v2.0.2 [https://palletsprojects.com/], Numpy v1.21.0 [19], Pandas v1.2.3 [20, 21], scipy v1.8.1 [22], psycopg2 v2.9.3 [https://www.psycopg.org/] and matplotlib v3.4.2 [23]. The data is stored in a PostgreSQL v12.10 [https://www.postgresql.org/] relational database. The frontend application is implemented using the Angular Framework v13.3.0 [https://angular.io/] including a SwaggerUI API documentation. The genome browser was realized using JBrowse 2 v2.1.4 [24]. Frontend and backend applications are dockerized through Docker v25.0.4 [25]. The source code and the dockerized database can be found at: https://gitlab.gwdg.de/MedBioinf/mtb/cnv-database.

### Data resources and processing

For CNV in cancer data, we primarily used the COSMIC dataset, specifically the file *Cosmic_CompleteCNA_Tsv_v97_GRCh38*.*tar* [26]. For cancer cell line data, we used the *CCLE_gene_cn*.*csv* file from the DepMap portal (https://depmap.org/portal), release 22Q2 [27]. As a negative control, we included CNV data from the Database of Genomic Variants (DGV) and dbVAR [28, 29].

For the cancer patient in the PanCNV-Explorer, we utilized the COSMIC dataset, which contains pre-normalized data from the International Cancer Genome Consortium (ICGC) and The Cancer Genome Atlas (TCGA) programs [26]. The genes located in the regions of CNVs were examined and mapped using SeqCAT to obtain the necessary gene information [30]. For cell line cancer data, we collected information from the DepMap portal [27] and classified CNV segmentation means, mapping those below 0.7 as gene deletions and those above 1.3 as gene amplifications. Rather than using precise CNV ranges, we assigned start and stop positions for each gene based on its CNV status. Therefore we encounter a high amount pseudo-CNV for the cell line data. In addition, we mapped cancer types from cell lines to their corresponding counterparts in the COSMIC dataset to ensure a consistent data resource. To identify non-cancer variations, we used CNV data from DGV and dbVAR as a control set representing the healthy population [28, 29]. All data were processed according to the described methods and stored in a PostgreSQL database, accessible via a Python Flask API.

## Results

### Data processing

Several normalisation steps are performed in PanCNV-Explorer to make the data consistent and comparable, so that other researchers can use the data directly in their own pipelines or for their own research questions. For cancer patients, pre-normalised data from COSMIC were used and mapped to the underlying genes. For cancer cell lines, data from DepMap were classified as gene deletions or amplifications based on CNV segmentation means. Instead of precise CNV ranges, genes were assigned start and stop positions based on CNV status. Cancer entities from the cell lines were manually matched to patient data to ensure consistency. Non-cancer variations were identified using control data from healthy population sources such as dbVAR and DGV. All data were processed and stored in a database accessible via an API.

### Comprehensive data resource

The dataset contains a total of 5,482,592 CNVs derived from 15,809 samples. They are divided into deleterious and benign data. The deleterious data were imported from ClinVar for patient data and DepMap for cell line data. ClinVar contributed 957,677 CNVs, while DepMap contributed 4,155,909 CNVs. The large number in the DepMap dataset is due to the method of mapping. In total the dataset contains CNVs from 36 different tissues and includes 21 primary cancer types, excluding subtypes.

In contrast, the benign data were obtained from the Database of Genomic Variants (DGV) and dbVar. DGV contributed 342,447 CNVs, while dbVar added 26,559 CNVs to the dataset. This collection of deleterious and benign CNVs allows a comprehensive analysis across different tissues and cancer types.

### Analysis and visualization of CNVs in cancer entities

For each cancer type, we quantified the number of affected genes to distinguish between amplifications and deletions. Based on this, a p-value was calculated for each gene within each cancer type to assess whether a gene was significantly more affected compared to the same gene in other cancer types. The p-value was calculated using Fisher’s exact test. All CNVs from the benign dataset were mapped to their corresponding genes and annotated to indicate whether the same genes were also found in the cancer dataset.

We continued to analyze the full genomic spans of CNVs by assessing whether each position in the human genome was involved in a copy number event. Amplifications and deletions were considered separately, and chromosome-specific plots were generated for each cancer type to visualise the affected regions. These plots (see Figure 1, 2) were then extended to compare two cancer types, allowing us to identify regions that are commonly affected across cancers as well as those that are unique to specific cancer types.

**Figure 1:**
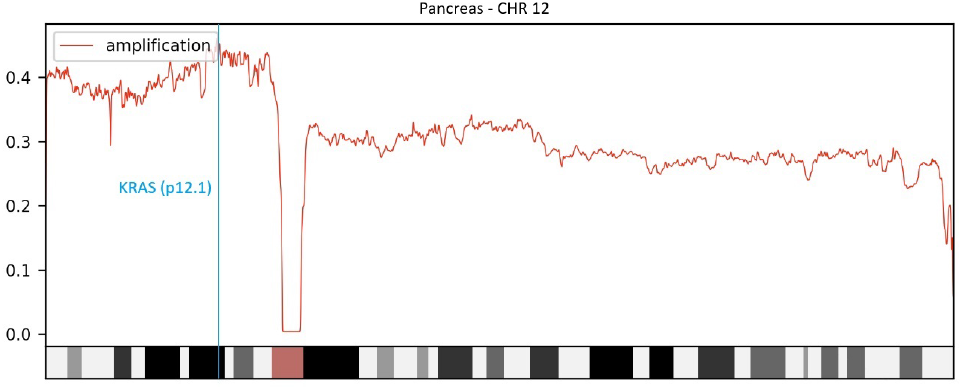
Amplification of chromosome 12 in pancreatic cancer. Amplification of chromosome 12 is a common finding in pancreatic cancer. Specifically, the short arm (p-arm) shows amplification in about 40% of patients, while the long arm shows a notable amplification frequency of about 30%. Of particular interest is the amplification of the KRAS gene (blue line) on the p-arm, which occurs in more than 40% of patients. KRAS is a well-established oncogene that is frequently involved in pancreatic cancer.

**Figure 2:**
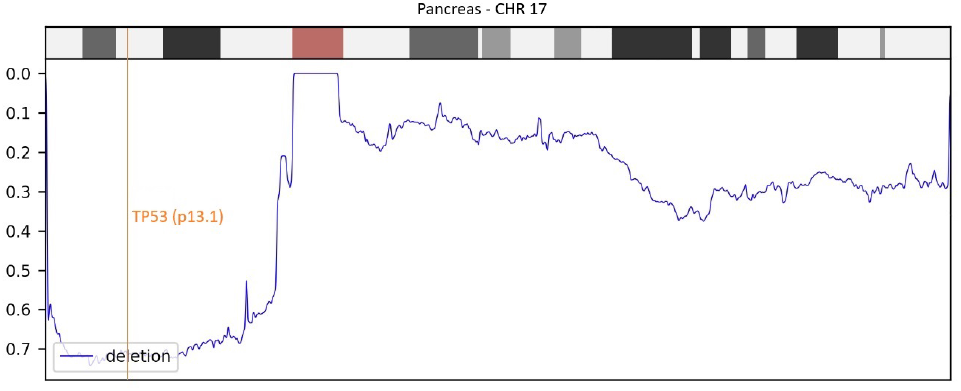
Deletions on chromosome 17 in pancreatic cancer. Deletions on chromosome 17 are common in pancreatic cancer, particularly on the short arm (p-arm), where deletions occur in about 70% of patients. The long arm shows a lower frequency, starting at about 20% and increasing to about 30% overall. In particular, deletions of the TP53 gene (orange line) on the p-arm occur in about 70% of patients. TP53 is a well-known tumour suppressor gene that is often associated with cancer as a hallmark of the disease.

The generated plots highlight a significant frequency of CNV amplifications in pancreatic cancer, particularly on chromosome 12, where approximately 40% of the samples show amplification. This is of notable interest because the KRAS oncogene is located in this region (Figure 1), this number is also frequently reported in other studies for around 35%-45% this entity [31, 32]. In addition, analysis of the short arm of chromosome 17 reveals a high deletion frequency, affecting approximately 70% of samples (Figure 2). This region contains the tumor suppressor gene TP53, another well-known gene frequently altered in pancreatic cancer [33].

### Use Case 1: Compare two cancers for similarities and differences

A typical research question in pancreatic cancer is the role of the MYC gene in this disease. It is therefore of interest to determine whether MYC is specific to pancreatic cancer or whether it plays a more general role in other types of cancer. Using the PanCNV-Explorer allows the visualisation of CNV plots across different cancer types and the comparison of two cancer entities in parallel. This feature facilitates the identification of genomic regions affected by CNVs in both cancers, highlights genes that are commonly altered in certain cancers, and shows areas that are uniquely affected. In addition, the database provides a table view that lists the most affected genes for the selected cancer entity and chromosome, further supporting the comparative analysis of CNVs across cancers. In Figure 3, a comparison of CNV patterns in breast and pancreatic cancers on chromosome 8 are shown. Notably, differences are observed on the short arm of the chromosome, where CNV amplifications occur in approximately 0.05% of breast cancer cases, compared to about 0.2% in pancreatic cancer. On the long arm, the CNV frequencies are more similar, especially in the second half of the arm. One significant feature in this region is the MYC gene, a well-known oncogene. Activation of MYC is a hallmark of cancer and is considered one of the most potent drivers of tumorigenesis [34]. In our analysis, MYC amplifications were detected at high frequencies in both breast cancer (45%) and pancreatic cancer (55%). This leads to the conclusion that MYC amplification seems to be one of the main causes in the development of breast and pancreatic cancer. By comparing pancreatic cancer with other cancers, similar patterns can also be seen for ovarian, liver and eye cancers.

**Figure 3:**
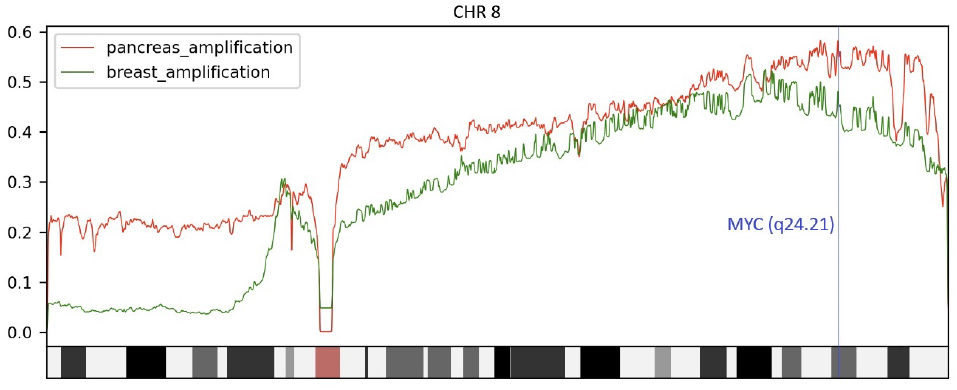
Comparison of amplifications on chromosome 8 in pancreatic and breast cancer. Amplification on chromosome 8 differs significantly between pancreatic and breast cancer. On the short arm of the chromosome, breast cancer has a general amplification frequency of around 6%, while pancreatic cancer has a significantly higher frequency of around 22%. Near the centromere, both cancers show a peak in amplification frequency, rising to about 30%. On the long arm of the chromosome, the second half shows similar amplification frequencies in both cancers, ranging from 40% to 50%. This region contains the MYC oncogene (blue line), which is known to be a hallmark of cancer.

### Use Case 2: Genome Browser to identify commonly affected genes

In another pancreatic cancer study, the PanCNV-Explorer identified highly frequent deletions in the q-arm of chromosome 16. An important question is whether these deletions are by chance or overlap with similar defects in other cancers, including the involvement of potential key genes. By comparing this finding with breast cancer, another region with a high deletion frequency can be found. The next step is to find out which genes in this region overlap between the two cancers. As shown in Figure 4, frequent CNV deletions in chromosome 16 are also prominent in breast cancer, consistent with one of the most common genetic alterations observed in this cancer [35]. This observation suggests the potential involvement of cancer driver genes such as *TRADD, DDX19, PSKH1, WWP2, PARD6A* and *BCAR1* in disease progression. The integrated genome browser allows further exploration of this region to identify additional genes that may also be associated with pancreatic cancer.

**Figure 4:**
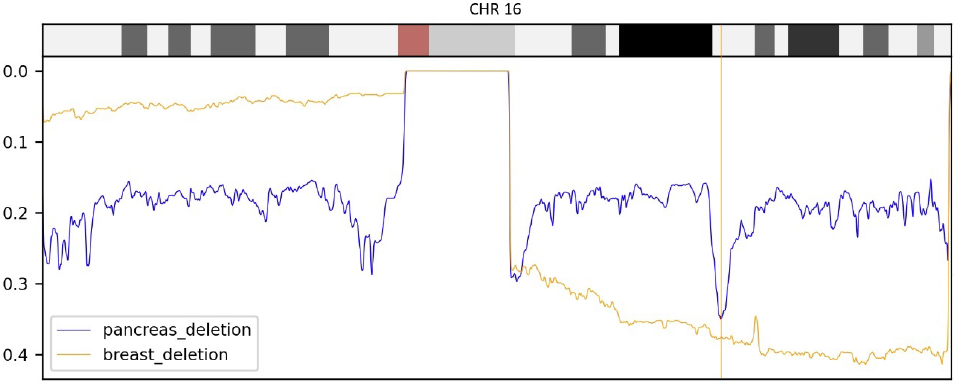
Comparison of deletions on chromosome 16 in pancreatic and breast cancer. Deletions on chromosome 16 show different patterns between pancreatic and breast cancer. On the short arm, breast cancer has a general deletion frequency of about 8%, whereas pancreatic cancer has a higher frequency of 20% to 25%. On the long arm, pancreatic cancer and breast cancer show different frequencies: about 20% in pancreatic cancer compared to 30% to 40% in breast cancer. A notable peak in pancreatic cancer shows deletion frequencies rising to around 35%, highlighting an interesting region (indicated by the orange vertical line). The literature identifies genes such as *TRADD, PARD6A* and *PSKH1* as potential tumour suppressor genes in breast cancer, and these genes also appear to be more frequently affected in pancreatic cancer.

Figure 5 presents a zoomed-in view of the peak previously identified in Figure 4. By displaying the gene tracks along chromosome 16, we can observe all the genes located within the region of high deletion frequency. In particular, three genes *TNFRSF1A* Associated Via Death Domain (*TRADD*), partitioning defective 6 homolog alpha (*PARD6A*), and protein serine kinase H1 (*PSKH1*) are highlighted, which have been described as potential tumor suppressors in breast cancer [35]. *TRADD* is involved in *TNF* receptor signaling and apoptosis [36], *PARD6A* plays a key role in cell polarity and asymmetric cell division [37], and *PSKH1* functions in pre-mRNA splicing and may contribute to cellular signaling pathways [38]. In addition, dipeptidase-2 (*DPEP2*), a member of the *DPEP* family known for dipeptide hydrolysis and found to be underexpressed in lung adenocarcinoma [39], also emerges as an interesting candidate. The deletion of these prognostic genes in pancreatic cancer suggests their potential relevance beyond breast and lung cancer. This demonstrates the importance of this database in pan-cancer research to identify common mutated genes along different cancer entities.

**Figure 5:**
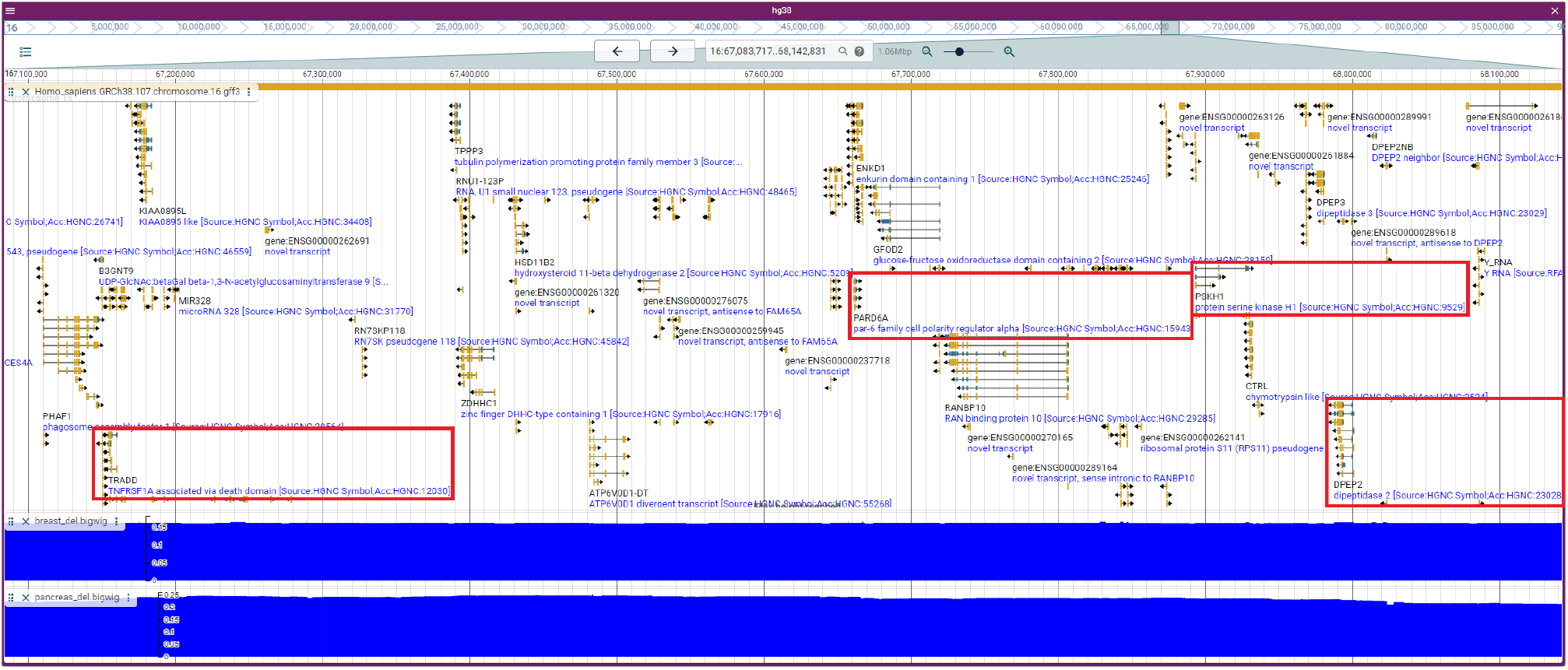
Genome browser view of chromosome 16, focusing on the peak position Figure 4. The Genome Browser provides detailed information about the genes located at the identified peak position. By comparing these genes with the published literature, we identified several potential tumour suppressor genes (TSGs) for breast cancer, including *TRADD, PARD6A, PSKH1* and *DPEP2* (highlighted in red boxes). In addition, both pancreatic and breast cancers show a high frequency of deletions in this region.

## Discussion

In summary, the PanCNV-Explorer represents a valuable and comprehensive resource for a pan-cancer analysis of copy number variations across multiple cancer types. The database integrates data from established repositories, such as COSMIC and dbVAR [26, 29], and provides tools for detailed exploration of CNV data, including amplification and deletion events across multiple genomic regions. Through a user-friendly interface, researchers can easily access visualizations and statistical analyses that provide insight into the distribution of CNVs and their potential role in tumorigenesis.

We highlight potential use cases for the PanCNV-Explorer, such as the detection of recurrent CNV events in specific cancers, including significant amplifications in oncogenes, like *MYC* [7, 40] and *KRAS* [41], and how new genes can be discovered by comparing different entries. This was demonstrated in the second use case, where known cancer-related genes from breast cancer, such as *TRADD, PARD6A* and *PSKH1* [35], were linked to pancreatic cancer. The PanCNV-Explorer is thus opening up new possibilities for cancer research by facilitating the identification of potential cross-cancer genetic associations.

PanCNV-Explorer currently offers a range of analysis methods, with the potential for future expansion to include additional techniques required by ongoing research projects. While these methods are not yet included, we provide an API for data download to enable researchers to implement their own custom analysis pipelines. One possible extension could be to investigate the relationship between SNVs and specific CNVs. In addition, the inclusion of a broader data set would facilitate the analysis of cancer subtypes as distinct entities. For example, this could allow more refined studies of gliomas and meningiomas in the central nervous system tumours [42] and mature T-cell, NK-cell and B-cell neoplasms in lymphomas [43] or other cancer entities.

The database’s ability to quantify and compare CNVs across cancer types is particularly important for understanding tumor heterogeneity. In summary, the PanCNV-Explorer is a valuable pan-cancer resource for researchers focused on genomics, providing a robust and accessible platform for exploring CNVs across a broad spectrum of cancer types. Its ease of use and comprehensive data make it an valuable tool for further cancer research.

## Data Availability

All data produced are available online at: https://gitlab.gwdg.de/MedBioinf/mtb/cnv-database

https://cancer.sanger.ac.uk/cosmic

https://depmap.org/portal/

https://dgv.tcag.ca/dgv/app/home

https://www.ncbi.nlm.nih.gov/dbvar/

## Author Contributions

Idea and design: KK, JD. Backend, frontend and database implementation: KK. Data harmonization, annotation and analysis: KK. Paper writing: KK JD.

## References

[1] Richard Redon et al. “Global Variation in Copy Number in the Human Genome”. In: Nature 444.7118 (Nov. 2006), pp. 444–454. issn: 0028-0836, 1476-4687. doi: 10.1038/nature05329. (Visited on 09/13/2024).

[2] Xin Shao et al. “Copy Number Variation Is Highly Correlated with Differential Gene Expression: A Pan-Cancer Study”. In: BMC Med Genet 20.1 (Dec. 2019), p. 175. issn: 1471-2350. doi: 10.1186/s12881-019-0909-5. (Visited on 09/13/2024).

[3] Feng Zhang et al. “Copy Number Variation in Human Health, Disease, and Evolution”. In: Annu. Rev. Genom. Hum. Genet. 10.1 (Sept. 2009), pp. 451–481. issn: 1527-8204, 1545-293X. doi: 10.1146/annurev.genom.9.081307.164217. (Visited on 09/21/2024).

[4] Mehdi Zarrei et al. “A Copy Number Variation Map of the Human Genome”. In: Nat Rev Genet 16.3 (Mar. 2015), pp. 172–183. issn: 1471-0056, 1471-0064. doi: 10.1038/nrg3871. (Visited on 09/13/2024).

[5] Alan M. Rice and Aoife McLysaght. “Dosage Sensitivity Is a Major Determinant of Human Copy Number Variant Pathogenicity”. In: Nat Commun 8.1 (Feb. 2017), p. 14366. issn: 2041-1723. doi: 10.1038/ncomms14366. (Visited on 09/21/2024).

[6] Adam Shlien and David Malkin. “Copy Number Variations and Cancer”. In: Genome Med 1.6 (2009), p. 62. issn: 1756-994X. doi: 10.1186/gm62. (Visited on 09/21/2024).

[7] Renumathy Dhanasekaran et al. “The MYC Oncogene — the Grand Orchestrator of Cancer Growth and Immune Evasion”. In: Nat Rev Clin Oncol 19.1 (Jan. 2022), pp. 23–36. issn: 1759-4774, 1759-4782. doi: 10.1038/s41571-021-00549-2. (Visited on 09/21/2024).

[8] Jana Hof et al. “Mutations and Deletions of the TP53 Gene Predict Nonresponse to Treatment and Poor Outcome in First Relapse of Childhood Acute Lymphoblastic Leukemia”. In: JCO 29.23 (Aug. 2011), pp. 3185–3193. issn: 0732-183X, 1527-7755. doi: 10.1200/JCO.2011.34.8144. (Visited on 09/21/2024).

[9] Chenhao Zhou et al. “Integrated Analysis of Copy Number Variations and Gene Expression Profiling in Hepatocellular Carcinoma”. In: Sci Rep 7.1 (Sept. 2017), p. 10570. issn: 2045-2322. doi: 10.1038/s41598-017-11029-y. (Visited on 09/21/2024).

[10] Kangpeng Yan et al. “Copy Number Variants Landscape of Multiple Cancers and Clinical Applications Based on NGS Gene Panel”. In: Annals of Medicine 55.2 (Dec. 2023), p. 2280708. issn: 0785-3890, 1365-2060. doi: 10.1080/07853890.2023.2280708. (Visited on 09/21/2024).

[11] Ana R. Cardoso et al. “Major Influence of Repetitive Elements on Disease-Associated Copy Number Variants (CNVs)”. In: Hum Genomics 10.1 (Dec. 2016), p. 30. issn: 1479-7364. doi: 10.1186/s40246-016-0088-9. (Visited on 09/21/2024).

[12] Malgorzata Bzymek and Susan T. Lovett. “Instability of Repetitive DNA Sequences: The Role of Replication in Multiple Mechanisms”. In: Proc. Natl. Acad. Sci. U.S.A. 98.15 (July 2001), pp. 8319–8325. issn: 0027-8424, 1091-6490. doi: 10.1073/pnas.111008398. (Visited on 09/21/2024).

[13] Stephanie B. Greene et al. “Chromosomal Instability Estimation Based on Next Generation Sequencing and Single Cell Genome Wide Copy Number Variation Analysis”. In: PLoS ONE 11.11 (Nov. 2016). Ed. by Aamir Ahmed, e0165089. issn: 1932-6203. doi: 10.1371/journal.pone.0165089. (Visited on 09/21/2024).

[14] Samuel Doré et al. “Exploring the Prognostic Significance of Arm-Level Copy Number Alterations in Triple-Negative Breast Cancer”. In: Oncogene 43.26 (June 2024), pp. 2015–2024. issn: 0950-9232, 1476-5594. doi: 10.1038/s41388-024-03051-y. (Visited on 09/13/2024).

[15] Daisy J. A. Oketch, Matteo Giulietti, and Francesco Piva. “Copy Number Variations in Pancreatic Cancer: From Biological Significance to Clinical Utility”. In: IJMS 25.1 (Dec. 2023), p. 391. issn: 1422-0067. doi: 10.3390/ijms25010391. (Visited on 09/13/2024).

[16] Luciano Cascione et al. “DNA Copy Number Changes in Diffuse Large B Cell Lymphomas”. In: Front. Oncol. 10 (Dec. 2020), p. 584095. issn: 2234-943X. doi: 10.3389/fonc.2020.584095. (Visited on 09/21/2024).

[17] Chun-Ting Lee, William J. Freed, and Deborah C. Mash. “CNVs in Neurodevelopmental Disorders”. In: Oncotarget 6.21 (July 2015), pp. 18238– 18239. issn: 1949-2553. doi: 10.18632/oncotarget.4853. (Visited on 09/21/2024).

[18] Amrita Chattopadhyay et al. “CNVIntegrate: The First Multi-Ethnic Database for Identifying Copy Number Variations Associated with Cancer”. In: Database 2021 (July 2021), baab044. issn: 1758-0463. doi: 10.1093/database/baab044. (Visited on 09/13/2024).

[19] Charles R. Harris et al. “Array Programming with NumPy”. In: Nature 585.7825 (Sept. 2020), pp. 357–362. issn: 0028-0836, 1476-4687. doi: 10.1038/s41586-020-2649-2. (Visited on 09/21/2024).

[20] The pandas development team. Pandas-Dev/Pandas: Pandas. Zenodo. Sept. 2024. doi: 10.5281/ZENODO.3509134. (Visited on 09/21/2024).

[21] Wes McKinney. “Data Structures for Statistical Computing in Python”. In: Python in Science Conference. Austin, Texas, 2010, pp. 56–61. doi: 10.25080/Majora-92bf1922-00a. (Visited on 09/21/2024).

[22] Pauli Virtanen et al. “SciPy 1.0: Fundamental Algorithms for Scientific Computing in Python”. In: Nat Methods 17.3 (Mar. 2020), pp. 261–272. issn: 1548-7091, 1548-7105. doi: 10.1038/s41592-019-0686-2. (Visited on 09/21/2024).

[23] John D. Hunter. “Matplotlib: A 2D Graphics Environment”. In: Comput. Sci. Eng. 9.3 (2007), pp. 90–95. issn: 1521-9615. doi: 10.1109/MCSE.2007.55. (Visited on 09/21/2024).

[24] Colin Diesh et al. “JBrowse 2: A Modular Genome Browser with Views of Synteny and Structural Variation”. In: Genome Biol 24.1 (Apr. 2023), p. 74. issn: 1474-760X. doi: 10.1186/s13059-023-02914-z. (Visited on 09/21/2024).

[25] Dirk Merkel. “Docker: lightweight Linux containers for consistent development and deployment”. In: Linux J. 2014.239 (Mar. 2014). issn: 1075-3583.

[26] Zbyslaw Sondka et al. “COSMIC: A Curated Database of Somatic Variants and Clinical Data for Cancer”. In: Nucleic Acids Research 52.D1 (Jan. 2024), pp. D1210–D1217. issn: 0305-1048, 1362-4962. doi: 10.1093/nar/gkad986. (Visited on 09/13/2024).

[27] Broad DepMap. DepMap 23Q4 Public. 2023. doi: 10.25452/FIGSHARE.PLUS.24667905.V2. (Visited on 09/13/2024).

[28] Jeffrey R. MacDonald et al. “The Database of Genomic Variants: A Curated Collection of Structural Variation in the Human Genome”. In: Nucl. Acids Res. 42.D1 (Jan. 2014), pp. D986– D992. issn: 0305-1048, 1362-4962. doi: 10.1093/nar/gkt958. (Visited on 09/13/2024).

[29] Ilkka Lappalainen et al. “dbVar and DGVa: Public Archives for Genomic Structural Variation”. In: Nucleic Acids Research 41.D1 (Nov. 2012), pp. D936–D941. issn: 0305-1048, 1362-4962. doi: 10.1093/nar/gks1213. (Visited on 09/21/2024).

[30] Kevin Kornrumpf et al. “SeqCAT: Sequence Conversion and Analysis Toolbox”. In: Nucleic Acids Research 52.W1 (July 2024), W116–W120. issn: 0305-1048, 1362-4962. doi: 10.1093/nar/gkae422. (Visited on 09/21/2024).

[31] Alyssa M Krasinskas et al. “KRAS Mutant Allele-Specific Imbalance Is Associated with Worse Prognosis in Pancreatic Cancer and Progression to Undifferentiated Carcinoma of the Pancreas”. In: Modern Pathology 26.10 (Oct. 2013), pp. 1346–1354. issn: 08933952. doi: 10.1038/modpathol.2013.71. (Visited on 09/21/2024).

[32] KhawlaS Al-Kuraya. “KRAS and TP53 Mutations in Colorectal Carcinoma”. In: Saudi J Gastroenterol 15.4 (2009), p. 217. issn: 1319-3767. doi: 10.4103/1319-3767.56087. (Visited on 09/21/2024).

[33] Jennifer P. Morton et al. “Mutant P53 Drives Metastasis and Overcomes Growth Arrest/Senescence in Pancreatic Cancer”. In: Proc. Natl. Acad. Sci. U.S.A. 107.1 (Jan. 2010), pp. 246– 251. issn: 0027-8424, 1091-6490. doi: 10.1073/pnas.0908428107. (Visited on 09/21/2024).

[34] M. Gabay, Y. Li, and D. W. Felsher. “MYC Activation Is a Hallmark of Cancer Initiation and Maintenance”. In: Cold Spring Harbor Perspectives in Medicine 4.6 (June 2014), a014241–a014241. issn: 2157-1422. doi: 10.1101/cshperspect.a014241. (Visited on 09/21/2024).

[35] Emad A. Rakha et al. “Chromosome 16 Tumor-suppressor Genes in Breast Cancer”. In: Genes Chromosomes & Cancer 45.6 (June 2006), pp. 527– 535. issn: 1045-2257, 1098-2264. doi: 10.1002/gcc.20318. (Visited on 09/21/2024).

[36] Yelena L. Pobezinskaya and Zhenggang Liu. “The Role of TRADD in Death Receptor Signaling”. In: Cell Cycle 11.5 (Mar. 2012), pp. 871–876. issn: 1538-4101, 1551-4005. doi: 10.4161/cc.11.5.19300. (Visited on 09/21/2024).

[37] Ziwen Lu et al. “Partitioning Defective 6 Homolog Alpha (PARD6A) Promotes Epithelial– Mesenchymal Transition via Integrin B1-ILKSNAIL1 Pathway in Ovarian Cancer”. In: Cell Death Dis 13.4 (Apr. 2022), p. 304. issn: 2041-4889. doi: 10.1038/s41419-022-04756-2. (Visited on 09/21/2024).

[38] G. Brede. “PSKH1, a Novel Splice Factor Compartment-Associated Serine Kinase”. In: Nucleic Acids Research 30.23 (Dec. 2002), pp. 5301–5309. issn: 13624962. doi: 10.1093/nar/gkf648. (Visited on 09/21/2024).

[39] Yuanyi Wang et al. “Dipeptidase-2 Is a Prognostic Marker in Lung Adenocarcinoma That Is Correlated with Its Sensitivity to Cisplatin”. In: Oncol Rep 50.2 (July 2023), p. 161. issn: 1021-335X, 1791-2431. doi: 10.3892/or.2023.8598. (Visited on 09/21/2024).

[40] Alexander Brik et al. “Digital PCR for the Analysis of MYC Copy Number Variation in Lung Cancer”. In: Disease Markers 2020 (Sept. 2020), pp. 1–8. issn: 0278-0240, 1875-8630. doi: 10.1155/2020/4176376. (Visited on 09/21/2024).

[41] Dae-Ho Choi et al. “Prevalence of KRAS Amplification in Patients with Metastatic Cancer: Real-world next-Generation Sequencing Analysis”. In: Pathology-Research and Practice 261 (Sept. 2024), p. 155473. issn: 03440338. doi: 10.1016/j.prp.2024.155473. (Visited on 09/21/2024).

[42] David N Louis et al. “The 2021 WHO Classification of Tumors of the Central Nervous System: A Summary”. In: Neuro-Oncology 23.8 (Aug. 2021), pp. 1231–1251. issn: 1522-8517, 1523-5866. doi: 10.1093/neuonc/noab106. (Visited on 09/21/2024).

[43] Rita Alaggio et al. “The 5th Edition of the World Health Organization Classification of Haematolymphoid Tumours: Lymphoid Neoplasms”. In: Leukemia 36.7 (July 2022), pp. 1720– 1748. issn: 0887-6924, 1476-5551. doi: 10.1038/s41375-022-01620-2. (Visited on 09/11/2024).

